# Effectiveness of Focal Muscle Vibrations in Improving Sensorimotor Performance, Mobility and Strength in Spinal Cord Injury Population: A Systematic Review

**DOI:** 10.1101/2023.12.22.23300431

**Authors:** Moeez Ashfaque, Amit N. Pujari, Imran Khan Niazi, Imran Amjad, Heidi Haavik, Simon F. Farmer

## Abstract

**Introduction:** The pathophysiology of spinal cord injury (SCI) is still not completely understood. Current SCI rehabilitation strategies remain ineffective. Focal muscle vibrations (FMVs), through afferent nerve stimulation, modulate peripheral and central pathways and have potential as a complementary and easy to administer rehabilitation tool in treating SCI populations, however the exact effectiveness of FMVs remains unknown.

**Methods:** This study is a systematic review on the use of FMVs in SCIs. Sensorimotor function, and mobility and strength were the main outcomes considered. Science Direct, PubMed, Cochrane library, PEDro, Google Scholar and Springer databases were searched for original studies until June 2023. The PEDro scale was used to assess the methodological quality of the studies.

**Results:** Twenty-three studies were included. Nine studies using FMV in the upper limb and fourteen in the lower limb. The analysis includes 422 SCI patients and 132 non-disabled participants, with a focus on male, chronic SCI cases, and a prevalence in North American studies.

**Conclusion:** Our findings suggest potential benefits of FMV on sensory perception, motor function, mobility, and strength in both upper and lower limbs of SCI patients. However, there is acute need for further research to optimize the application of FMV through its vibration parameters, location, duration and understanding its long-term effects. Due to the lack larger sample sizes and longitudinal studies, any conclusions derived here and within these studies should be interpreted with caution.

WHAT IS ALREADY KNOWN ON THIS TOPIC
Focal muscle vibrations (FMVs) are known to modulate central and peripheral pathways by a the afferent nerves and have been shown to improve neuro-muscular performance in healthy, stroke and spinal cord injury (SCI) populations.

WHAT THIS STUDY ADDS
This study synthesises the current evidence to assess the effects of FMVs on the sensorimotor function and the rehabilitation of SCI subjects. Our study also highlights the limitations of current evidence.

HOW THIS STUDY MIGHT AFFECT RESEARCH, PRACTICE OR POLICY
The evidence presented and discussed here offers valuable insights into the effectiveness of FMV in SCI rehabilitation, its usefulness in improving functional movements, presents insights into the underlying neural mechanisms, and offers recommendations for both clinical applications and future research.

## 1. INTRODUCTION

A spinal cord injury (SCI) is a damage to the spinal cord, causing paralysis and sensory deficits below the injury level. It can be traumatic or non-traumatic, acute or chronic, paraplegic or tetraplegic, and complete or incomplete. It is estimated that 10.5 in 100,000 SCI cases emerged globally during 2006-2016 [29]. In the UK, each year 16 traumatic and 2-3 non-traumatic cases per million population are estimated [30]; in the US 40 per million traumatic cases are estimated [31]. SCI remains a profound and life-altering medical condition characterised by the disruption of sensory and motor pathways, often resulting in debilitating functional deficits [1]. Mobility, motor and sensory function loss, and strength decrease are among the major complications associated with the SCI [2]. Neurogenic bladder and bowel, urinary tract infections, pressure ulcers are also frequent complications [1]. These complications are directly related to the patient’s life expectancy and quality of life [1]. In addition to its life changing medical consequences SCI results in a financial burden on the patient, treatment bodies and the governments, with average estimated initial treatment cost at 142,366 USD and readmission costs over a decade at 49.4 million USD in the US [31]. Individual co-morbidities can account for up to 12900 USD per visit [31]. In the UK the overall average estimated lifetime treatment cost is estimated at 1.12 million GBP per patient [30]. In Bangladesh, a low to medium income country, 76% of patients were found pushed into illness-induced poverty [32]. There is a gender disbalance in its occurrence with around 80% being male patients [33]. SCI results in life-altering physical and sensory impairments, necessitating comprehensive care and rehabilitation [1]. The treatment landscape for SCIs encompasses a range of modalities, including surgical interventions, pharmacological therapies, and rehabilitative approaches [3, 4]. While surgical procedures aim to stabilise and repair the spinal cord, pharmacological treatments focus on managing symptoms like pain and spasticity. Rehabilitation plays a pivotal role in optimising function and quality of life for individuals with SCIs, encompassing physical therapy, occupational therapy, and assistive devices [1, 3, 4,34].

One emerging therapeutic modality that has received increasing interest in the past two decades is the application of vibrations, particularly focal muscle vibration (FMV), a non-invasive neuro-modulatory technique that may tap into the inherent potential of neuromuscular system for promoting functional recovery after neurological injury (such as SCI) or neurological disorders [5, 35,36, 37]. FMV involves the application of mechanical vibrations to specific muscle groups or tendons. These controlled vibrations alter transmission of primary and secondary muscle afferents (Ia, Ib, and type II afferents) [38, 39,40], cutaneous mechanoreceptors [46] and modulate cortical excitability [47, 48. FMV is gaining increasing interest in neurological disease management [5, 36]. It is being explored as an innovative primary and adjunctive therapy in various medical fields, including spinal cord injury rehabilitation to facilitate functional recovery and improve the overall quality of life for individuals with neurological impairments [5, 27, 41, 42, 43]. FMV offers a distinct advantage in SCI management by providing a safe and targeted approach to neuromodulation. Unlike invasive procedures, FMV does not require surgical intervention and it is easy to use, minimising associated risks. Pharmaceutical approaches (e.g. anti-spasticity agents) are typically non-targeted and are generally result in overall neural activity suppression [5] and possible side effects [5]. The ability of FMV to selectively target muscles and sensory receptors makes it a promising tool for enhancing muscle strength, reducing spasticity, and improving sensory perception, all of which are critical aspects of SCI recovery and rehabilitation. FMV’s non-invasive nature has a potential to make it a valuable complement to the existing treatment options for spinal cord injuries.

However, despite its potential benefits much of the research on the use FMV has been focussed on its use in stroke rehabilitation [44, 45]. As a result, utility and effectiveness of the FMV in SCI rehabilitation remains unclear. Therefore, this systematic review endeavours to explore and critically evaluate the existing body of literature ‘on the use of FMV to improve various aspects of spinal cord injury-related detriments’, particularly its effect on two critical areas of SCI recovery: (1) muscle strength and mobility, and (2) sensory and motor function in SCI patients. By synthesising the current evidence, this review aims to provide valuable insights into the potential efficacy and safety of FMV therapy in the management of spinal cord injury.

## 2. METHODS

### 2.1 Information sources

Preferred Reporting Items for Systematic Reviews and Meta-Analyses (PRISMA) [49] reporting methods were adopted. Science Direct, PubMed, Cochrane library, PEDro, Google Scholar and Springer databases were used to conduct the literature search.

### 2.2 Eligibility criteria and outcomes

Articles were screened that adhered to the following PICO scheme, Participants: SCI, Intervention: FMV, Comparison: absence of vibration, Outcome: sensory and motor function, or mobility and strength.

### 2.3 Search strategy

The keywords used for the search are provided in Table-1. Articles published until November 2023 were searched. The titles were screened initially, followed by abstracts and then full texts. Any theses/doctoral dissertations were not considered as it was not clear if they were peer reviewed. One review article was also removed for not being an original research article [5]. Further, if articles were investigating penile vibration, or sexual or related functions, they were also screened out. Notably, many articles fell into this category and may be suitable for a separate future systematic review. Only English texts were considered. Details of screening are outlined in Figure-1. Due to heterogeneity of the outcomes no meta-analysis was performed.

**TABLE 1.** List of the keywords used.

|  |
| --- |
| PEDro, Springer, Science Direct and Cochrane library |
| "spinal cord injury", muscle vibration |
| Google Scholar |
| "focal muscle vibration" OR "local muscle vibration" OR "segmental muscle vibration" OR "localized mechanical vibration" OR "focal tendon vibration" OR "muscle vibration" OR "Focal Vibro-Tactile Stimulation" OR "hand-arm vibration" OR "focal vibration" OR "proprioceptive stimulation" OR "repetitive sensory input" OR "vibration" OR "vibration stimulation" OR "local vibration" OR "localized vibration" AND "spinal cord injury" OR "Tetraplegia" OR "quadriplegia" OR "paraplegia" OR "spasticity" OR "spasm" OR "spastic paraplegia" OR "spastic paresis" OR "spinal cord lesion" OR "traumatic spinal lesion" OR "hypertonia" |
| PubMed |
| "Spinal Cord Injuries"[Mesh] OR "Quadriplegia"[Mesh] OR "Paraplegia"[Mesh] OR "spinal cord lesion"[tw] OR "traumatic spinal cord lesion" OR "Spasm"[Mesh] OR "spastic*"[tw] OR "hyperton*"[tw] OR "spastic paresis"[tw] AND "focal muscle vibration"[tw] OR "local muscle vibration"[tw] OR "segmental muscle vibration"[tw] OR "mechanical vibration"[tw] OR "tendon vibration"[tw] OR "muscle vibration"[tw] OR "focal vibration"[tw] OR "vibration"[tw] OR "vibration stimulation"[tw] OR "local vibration"[tw] OR "localized vibration"[tw] OR "Vibrotactile Stimulation"[tw] OR "hand-arm vibration"[tw] OR "proprioceptive stimulation"[tw] OR "repetitive sensory input"[tw]. |

**FIGURE-1:**
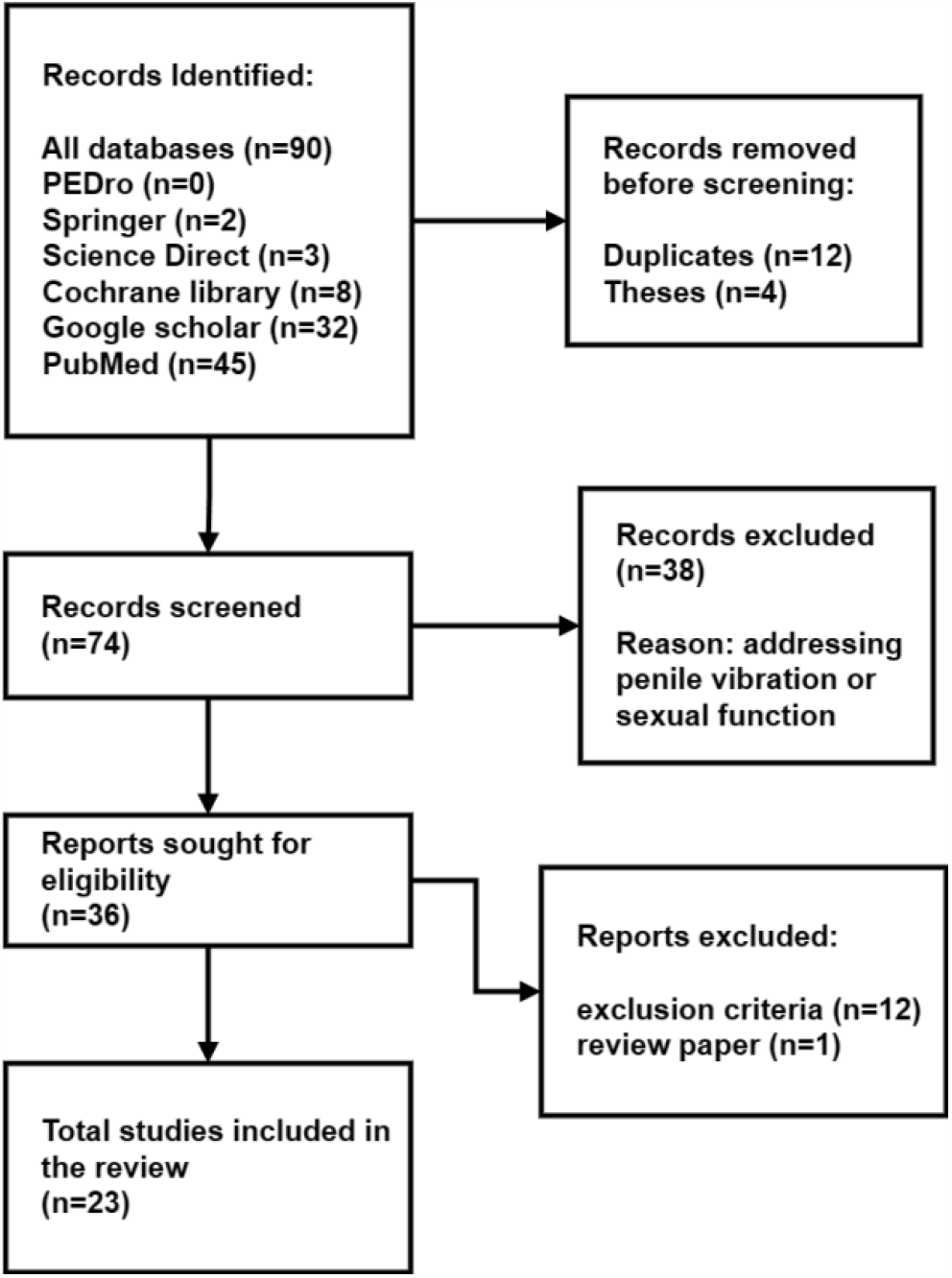
PRISMA Flow diagram. PRISMA flow diagram for papers identification and screening.

### 2.4 Data Synthesis

Article search and screening for eligibility was performed by one reviewer (MA) that adhered to the inclusion criteria. Data extraction was also performed by the same reviewer and included: author, year and country of the study, study design, outcome measures, participant demographics (mean age, gender), SCI characteristics (chronic/ acute, complete, incomplete, medications), sample size, limb of therapy and intervention elements (device, vibration frequency and amplitude, duration, sessions, muscle location and type). Any outstanding conflict was resolved by a second reviewer (ANP).

### 2.5 Methodological Quality Assessment

The PEDro scale was used to assess the methodological quality and the risk of bias of the included studies. One reviewer (MA) performed the methodological quality assessment. Studies were scored on the PEDro scale consisting of eleven items. One item on the PEDro scale (eligibility criteria) is related to external validity and is generally not used to calculate the method score, leaving a score range of 0–10 Please refer to table 4.

## 3. RESULTS

A total of 23 articles [6-28] were screened that qualified for the inclusion criteria. A total of 422 SCI patients (mean age: 38.36, gender: 324 male, 84 female, 14 not reported) and 138 non-disabled individuals (NDIs) (mean age: 33.01, gender: 73 male, 45 female, 14 not reported) took part in the FMV studies. In one study [16] multiple sclerosis and transverse myelitis patients were also part of the study, results of the study on SCI patients were only considered for this review. Remaining studies focused on SCI only. 264 patients were chronic, 27 were acute and the remaining 131 were not reported, 95 were complete and 174 were incomplete, while the remaining 154 were not reported to be either. In terms of countries, 14 studies are from North America (11 US, 3 Canada), 7 from Europe (2 Spain, 2 Italy and one each from Netherlands, France, and UK), one from Australia and one from East Asia (from Japan, published in 1996). Out of the 23 studies only 12 studies reported if an oral pharmaceutical medication was used or not. Of the reported people taking medications, baclofen was the most common (51 participants in total were reported administering it) followed by diazepam (8 participants, 5 of which were part of a study on diazepam [7]). Two participants used oxybutynin, one participant each was reported for clonazepam and dantrolene sodium. One study also reported the use of the following medicines (one participant each): gabapentin, furosemide, 4-aminopyridine and doxazosin. Fifteen different devices have been used to deliver FMVs, these devices are provided in Table 3. The methodological quality scores of the included studies ranged from 4-7, with fourteen studies scoring 5, one study scoring 4, four studies scoring 6, and two studies scoring 7. Details for each item are provided in Table 4. None of the studies fulfilled criterion 3 and 5 on the PEDro scale. Criteria 2,6 and 7 were also weak in many studies. Definitions of criteria are included in Table 4. Details specific to the upper limb (UL) and lower limb (LL) are listed below.

**TABLE 2:**
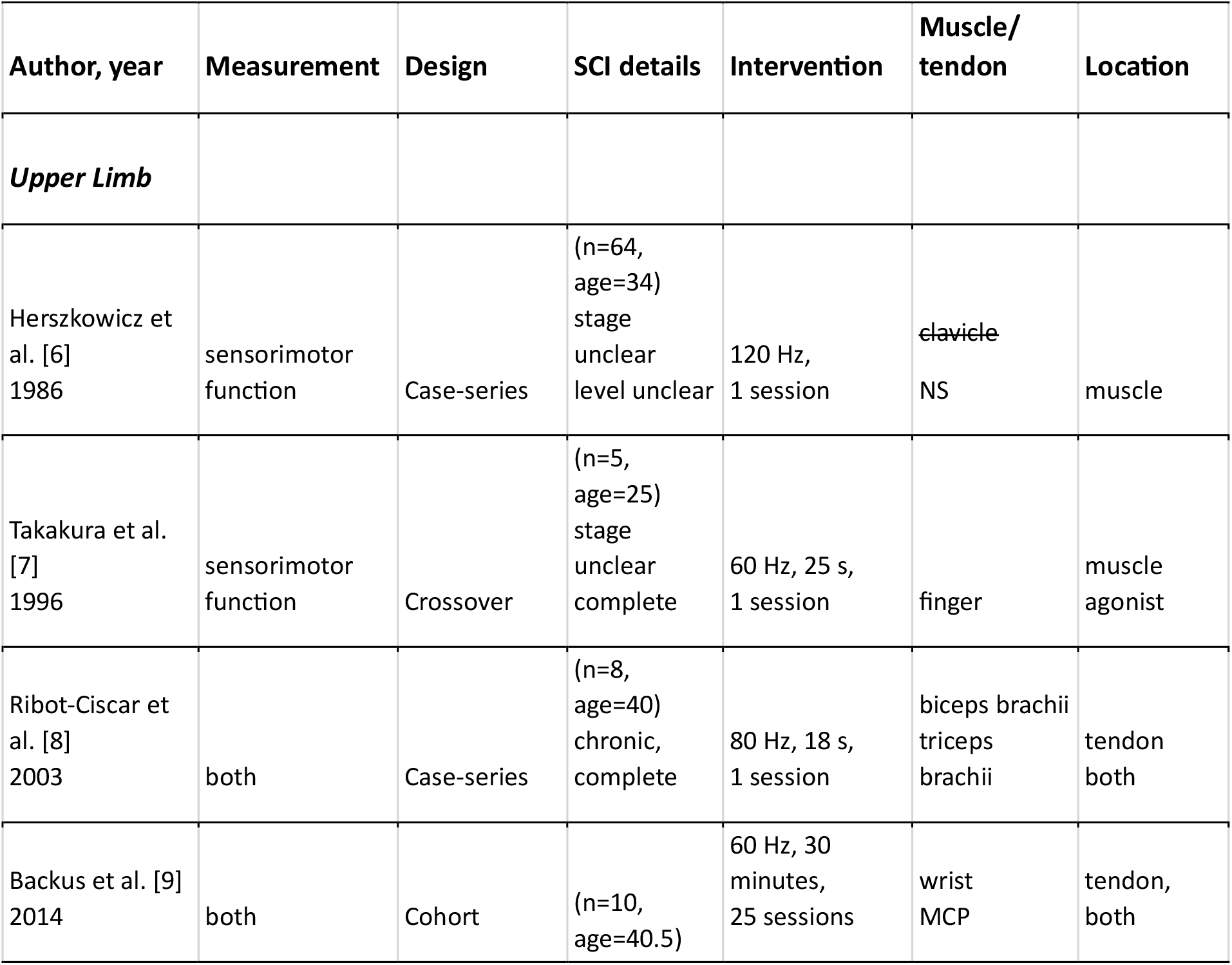
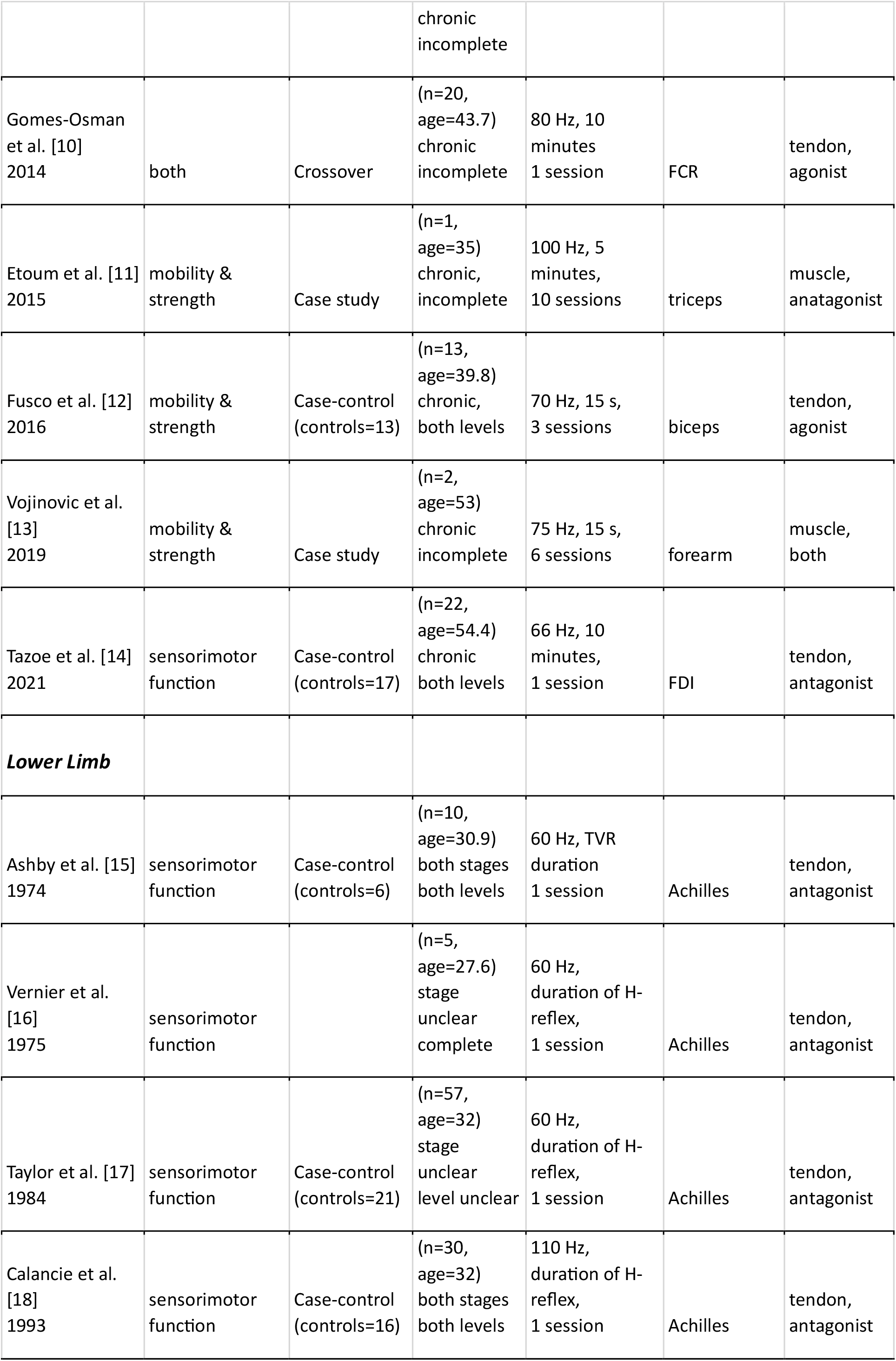
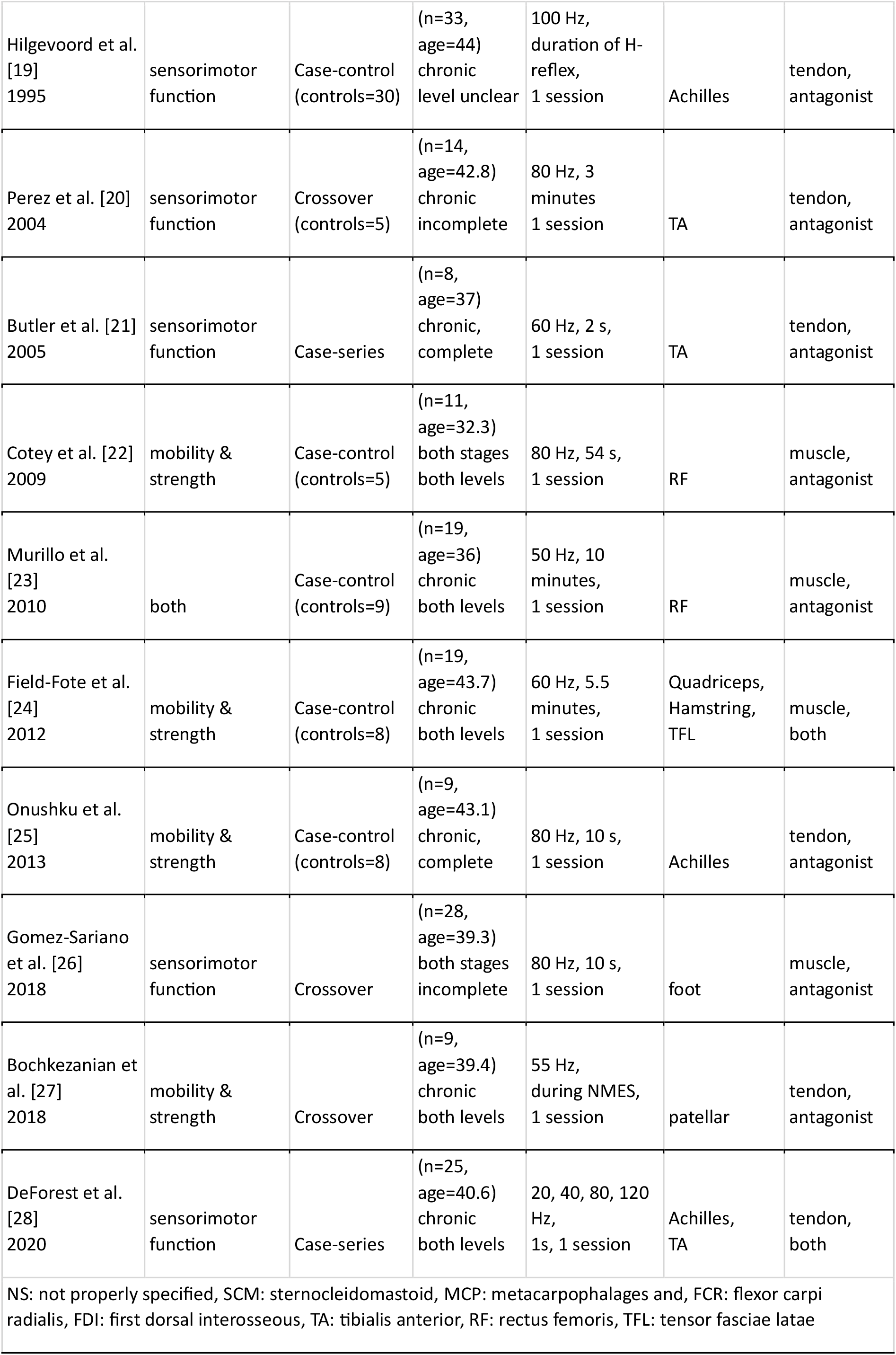
Characteristics of the studies included in the systematic review.

**TABLE 3:** List of the vibration devices used in the studies.

| Sr. | Device |
| --- | --- |
| 1 | Vibrameter [6] |
| 2 | Zeniter model TMT-18,<br>heiwa electronic industrial [8] |
| 3 | AMES technology [9] |
| 4 | CEN, USA [10] |
| 5 | Bosco system [11] |
| 6 | CroSystem nemoco [14] |
| 7 | Thrive no. 91 [18] |
| 8 | Breul and kjaer 4809 vibrator [19] |
| 9 | Heiwa denshi, model TNT-18 vibrator [20] |
| 10 | Pro massager USJ-301 [24] |
| 11 | Deep muscle stimulator [27] |
| 12 | Wahl jumbo vibrator[15, 17] |
| 13 | Wahl vibrator model 4196 sterling III [20] |
| 14 | Wahl powersage 4300 [23] |
| 15 | Custom made vibrators<br>[7,12,13,22,25,26,28] |
| 16 | Detail not provided [16] |

**TABLE 4:**
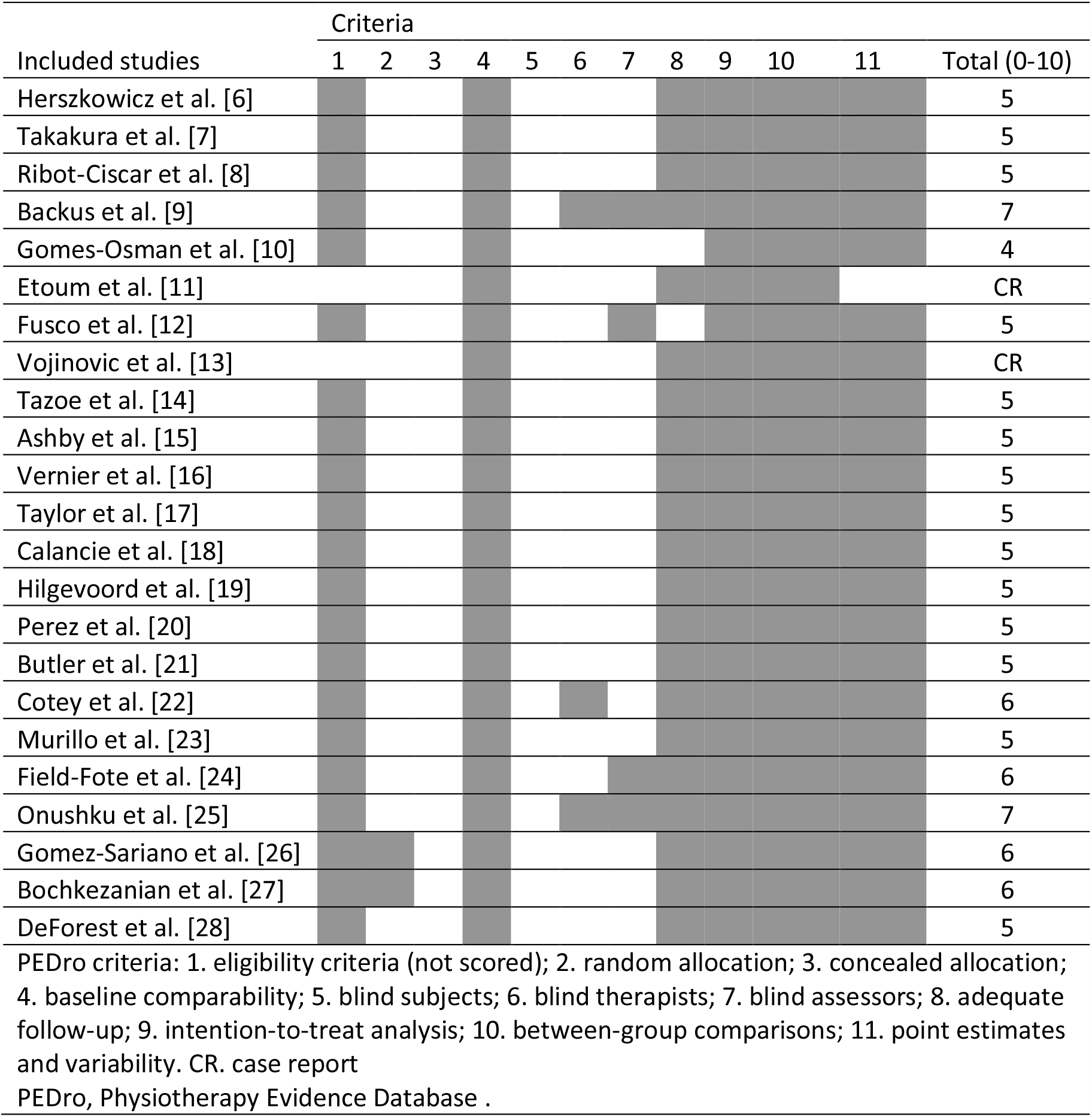
Methodological quality assessment of studies using the PEDro scale.

### 3.1 Use of FMV in the UL

Nine studies [6-14] addressed the application of FMV on the UL in SCI patients. Six of which focused on understanding the sensory and motor function in patients in response to FMV [6-10, 14] and six studies investigated the role of FMV in improving mobility and muscular strength in the UL [8-13]. Different outcome measures studied are: vibration sensation threshold [6], tonic vibration reflex (TVR) [8], corticomotor excitability [10] and cortical motor maps by transcranial magnetic stimulation (TMS) [14] were studied for assessing the effects on sensory and motor function, while for assessing mobility and strength effects, electromyography effects (EMG) [8], force [7-9], torque [9], range of motion (ROM) [9, 13], grip and release test (GRT) [9], 9 Hole Peg test [10], visuomotor tracking [10], modified Ashworth scale (MAS) [11,13] and questionnaires for mobility [9, 12, 13] were utilised. A total of 145 SCI participants were tested in the upper limb, with seven studies focusing on chronic SCIs (76 participants). In two studies (69 participants) it was not reported if the participants were chronic or acute. There was no study specifically targeting acute SCIs in the UL. Two studies were on complete SCIs (13 participants) and four studies on incomplete (33 participants). Two studies had both complete (8) and incomplete (27) participants, while one study did not mention the completeness details of the SCI (64 participants). Of the nine studies only two had NDIs as controls (30 participants) in addition to the SCI participants.

In terms of vibrational frequency for FMV, most of the studies fell within the 60-80 Hz range. Two studies used 60 Hz, one each for 66, 70 and 75 Hz, two for 80 Hz and one each for 100 Hz and 120 Hz. Only four studies reported the vibration amplitudes: two studies 2 mm, and one study each for 1 mm and 0.4 mm. Vibration was applied for less than half a minute in four studies (9 s, 15 s, 15 s and 25 s), 5 minutes in one study and 10 minutes in two studies. Two studies did not report the duration of application. Five studies had only a single session of vibration, and one each having 3, 6, 10 and 25 sessions. The following muscles were used for delivering FMVs: one study each for clavicle (skin of superior surface of the clavicle 8-10 cm from its sternal end), middle finger, wrist extensor, metacarpophalages (MCP) and first dorsal interosseous (FDI). One study applied to biceps brachii, one to triceps brachii and one applied on both biceps brachii and triceps brachii. One for flexor carpi radialis (FCR) while one for both extensor and flexor of the forearm. Of these, in five studies FMV was applied to tendons whereas in four studies it was applied to muscles. In four studies the muscle group was an agonist muscle, in one it was the antagonist, and in the other three studies both the antagonist and agonist muscles. Please refer to Table 2 for the details of the UL studies.

### 3.2 Use of FMV in the LL

Fourteen studies addressed the application of FMV on the LL [15-28]. Ten studies [15-21, 23, 26, 28] investigated the sensory and motor function while five [22-25, 27] studied the effect of FMV on mobility and strength in the lower limb. Outcome measures used to assess the function of the participants were: Hoffman reflex (H-reflex) [15-21, 26], TVR [15], tendon reflex [23] and EMG [21, 26, 28] for sensory and motor function, while EMG [22, 25], MAS [23], ROM [23] and torque [25, 27] were used for measuring mobility and strength. A total of 277 SCI participants were tested in the lower limb, with eight studies focusing only on chronic SCIs (136 participants) and in two studies (62 participants) it was unknown if the participants were chronic or acute. Four studies had both chronic and acute participants (79 individuals). There was no study specifically targeting acute SCIs in the LL. Four studies had both acute (27) and chronic (52) participants. Two studies were on complete SCIs (13 participants) and three studies on incomplete (51 participants). Seven studies had both complete (61) and incomplete (62 participants), while two studies did not mention the completeness of SCI (90 participants). Nine of the fourteen studies also had a NDI population (108 participants).

As far as vibrational frequency for FMV is concerned, 60 and 80 Hz were the most used. Five studies used 60 Hz and four used 80 Hz. One study each delivered 50 Hz, 55 Hz, 100 Hz and 110 Hz. One study tested four different vibration frequencies 20, 40, 80 and 120 Hz. Eight studies reported the vibration amplitudes: two 1 mm, and one each for 0.5mm, 1.5 mm, 2.2 mm, 3 mm, 4 mm, and 7 mm. In terms of duration of vibration, vibration was applied for the duration of obtaining the H-reflex in four studies [16-19], for the duration until a TVR was obtained in one [15] and for the duration of applying neuromuscular electrical stimulation (NMES) in one [27]. Vibration duration was less than 20 s in four studies: 1 s in [28] 2 s in [21], 10 s in [25] and 15 s in [26]. 60 s of vibrations were applied in [22], 3 minutes in [20], 5 minutes and 30 seconds in [24] and in [23] vibrations were applied for 10 minutes. All the LL studies were a single session study. Pertaining to the muscle of FMV application, six studies used the Achilles tendon, two studies delivered on tibialis anterior (TA) tendon, one study applied on both Achilles and TA tendon. Two studies used rectus femoris (RF) muscle, one study each for patellar tendon and plantar surface of foot, and in one study Quadriceps, hamstring, and tensor fasciae latae (TFL) muscles were used. Most of the studies were focusing on delivering FMV to tendons (10 studies) whereas in a few (4 studies) it was applied to muscles. In seven studies the muscle group was an antagonist muscle, in five studies it was the agonist, and in the other two studies both the antagonist and agonist muscles were used. Table 2 has details of the LL studies.

## 4. DISCUSSION

The results presented in this study provide valuable insights into the use of FMV as a potential therapeutic approach for individuals with SCI. The discussion will address key findings, implications, and limitations of the reviewed studies. It will also shed light on the current state of knowledge on the use of FMV for improving sensory and motor function and mobility and strength in the upper and lower limbs in SCI population.

The included studies involved a total of 422 SCI patients and 132 non-disabled (ND) participants. 76.77% of the SCI patients were male, reflecting the higher prevalence of research participation in SCI males, this is consistent with previous observations of higher occurrence rate (about 80%) in males [33]. Chronic SCI cases were more prevalent in the studies, indicating a focus on individuals with later stage injuries. Interestingly, the majority of the studies did not specify the completeness of the SCI, which could significantly impact the interpretation of the results. In regard to the geographical distribution of the research, majority of studies were conducted in North America, which may be indicative of availability of resources and/or regional differences in research interest and priorities. It is important to note that there was a significant focus on chronic SCI cases in both upper and lower limb studies, while the representation of acute SCI cases was limited. This imbalance in participant demographics could affect the generalizability of the findings to acute SCI populations. Future research should aim to include a more diverse range of SCI types and durations to better understand the potential benefits of FMV across different stages of the injury. The studies reviewed primarily utilised vibration frequencies in the range of 60-80 Hz, with 60 Hz and 80 Hz being the most common choices. This consistency in frequency selection may indicate an established optimal range for FMV interventions. However, the vibration amplitudes and durations varied across studies, which could further influence the effectiveness of FMV. As underlying neurophysiological mechanisms determining the effects of FMV in SCI are still poorly understand with only handful of studies providing this mechanistic evidence [28], further research is needed to determine the ideal parameters for FMV application in SCI rehabilitation.

### 4.1 Mobility and Strength

The use of FMV in ULs of SCIs present a mixed picture of its potential benefits on mobility. FMV applied to the biceps tendon in SCIs resulted in an illusion of arm movement, albeit smaller than in healthy individuals [12], suggesting potential for sensory perception in SCIs, though the extent is unclear. In contrast, a single ten-minute FMV session at the FCR tendon area during a functional task had no immediate impact on mobility measures [10]. The effectiveness of FMV may depend on session duration, frequency and stimulation location. Case studies revealed promise in long-term effects. Six FMV sessions (duration: 15 minutes each) in forearm muscles alongside functional tasks improved the MAS and ROM [13]. Even shorter (duration: 5 minutes each), 10-session FMV interventions in the triceps brachii showed lasting benefits (up to a month) in the MAS [11]. In a study involving 25 sessions of FMV in the hand region during functional tasks for 10 SCI patients, significant improvements were observed in grip and release performance as measured by the GRT and ROM [9]. GRT showed a 23% improvement immediately after the sessions, with a 7% further improvement three months post-treatment. In summary, FMV’s efficacy in SCIs is context-dependent, with potential for improvement in sensory perception and long-term benefits. The specific factors influencing its effectiveness, such as application location (tendon vs muscle), session duration, and combination with functional tasks require further exploration for optimal rehabilitation in SCIs.

FMV shows promise in improving upper limb strength in individuals with spinal cord injuries. However, its effects vary depending on factors such as muscle function and exposure duration. In complete SCIs, 9 seconds of FMV improved elbow extension force generation in the biceps brachii but was inconsistent in the triceps brachii, highlighting the differences in its effects on antagonist and agonist muscles [8]. Long term effects were not studied. A 10-minute session of FMV at the FCR tendon enhanced pinch force but only temporarily, the improvements did not last 30 minutes post the session [10]. And 25 sessions of FMV for 30 minutes to the antagonist hand muscles (metacarpophalangeal joint and wrist) in incomplete SCIs while performing a functional task enhanced the muscle strength [9]. Long term effects were again not studied. These findings suggest FMV’s potential in enhancing upper limb strength, but further research is needed to understand the impact of vibration location (tendon vs muscle, agonist vs antagonist), long term effects and differences in its effects on complete and incomplete pathologies.

FMV presents opportunities for improving lower limb muscle activity and gait dynamics in spinal cord injuries. However, nuances and unexplored aspects deserve attention. In one study [22], 54 seconds of FMV to the RF muscle boosted muscle activity in proximal quadriceps muscles during assisted gait but had no impact on distal leg muscles, suggesting the effects of FMV may be limited to stimulated muscle group(s) and its synergists/antagonists. In the same study, FMV also improved the transition between different phases of gait in SCI patients that had pathological gait. Another study [25] demonstrated that 10 seconds of Achilles tendon vibration during assisted hip movement increased muscle activity in the TA and GM muscles, particularly during hip flexion and with voluntary assistance. The effects of FMV with and without movement assistance, clinical outcomes, and long-term implications were however not studied in both these studies. A different study [23] extended FMV to 10 minutes on quadriceps, enhancing range of motion and MAS scores in SCIs, but it didn’t assess muscle activity and had varied outcome measures. Surprisingly, a brief 5.5 minutes of FMV to thigh muscles induced a step-like response in chronic SCIs [24], independent of SCI completeness and unaffected by locomotor training. The TFL muscle showed the most robust response. Furthermore, a study explored the combination of FMV and neuromuscular electrical stimulation [27], revealing improved muscular work capability in some SCIs, while others experienced decreased capability. The underlying mechanisms remain unclear. In conclusion, FMV holds promise for enhancing mobility in SCIs, but location specificity, movement assistance, clinical outcomes, and long-term effects need further investigation to optimise its use in rehabilitation.

### 4.2 Sensorimotor Function

The presented findings provide valuable insights into sensory perception and motor function in the ULs of SCIs. Notably, individuals with SCIs have a reduced sensory threshold at the clavicle, directly correlating with the injury level [6]. Furthermore, FMV interventions appear promising in restoring sensory perception, as demonstrated by significant improvements in finger digit sensation following a (30 minutes each) 25-session intervention [9]. 25 seconds of FMV to the middle finger resulted in a finger flexion reflex in complete SCIs, the amplitude of which was inhibited via acupuncture techniques [7]. SCI. Nine seconds of FMV always induces a TVR in the biceps brachii but only half the time in triceps brachii muscle [8]. Tonic vibration reflex (TVR) is a sustained contraction of a muscle after being subjected to vibration. Vibration excites muscle spindles, which in turn induce reflex contractions in the muscle being vibrated [8]. The inconsistent TVR response observed between the agonist and antagonist in upper arm muscles warrants investigation into its neurophysiological implications [8]. Additionally, the impact of vibration on motor function and neuroplasticity is evident, as 10 minutes of FCR tendon vibration increased long-term (30 minutes after intervention) corticomotor excitability [10]. A similar effect was observed with FMV at the FDI tendon, where motor maps generated by TMS expanded [14]. The immediate corticomotor excitability in [10] however did not change.

In the LL there is a larger number of studies investigating sensory and motor performance. The soleus H-reflex response to Achilles tendon vibration in the LL has been widely studied in literature and has offered valuable insights into neural mechanisms in the context of SCIs. Early studies by Ashby et al. [15] and subsequent validations [18, 19] have shown that, in the acute stage of SCI, vibration completely diminishes the soleus H-reflex, despite higher H-reflex amplitudes without vibration compared to NDIs. In the chronic stage, the H-reflex response ratio between vibration and no vibration (H_vib_/H) increases compared to NDI, indicating reduced H-reflex inhibition in response to vibration. [15, 18, 19]. Additionally, the TVR for the soleus muscle elicited by Achilles tendon vibration is completely or almost completely diminished in acute SCI, and this reduction persists in the chronic stage [15]. Diazepam’s influence on FMV action is also studied; the H_vib_/H ratio in complete SCIs does not change using Diazepam [16]. However, Diazepam’s effect on the H/M ratio is not known, which can provide further insights into the neural activity. Notably, as spasticity increases in SCI patients, the H_vib_/H ratio also rises [17], signifying reduced depression of the H reflex with vibration. The ratio increased with the duration of lesion for all patients and was unrelated to the level of lesion or the completeness of SCI [17]. Following neurophysiological implications are suggested by the author: a) mechanisms blocking the H-reflex become less effective as spasticity gains prominence, b) vibration produces greater background facilitation of motoneurons in spasticity conditions [17]. Calancie [18] noted that presynaptic inhibition is enhanced in acute SCI, contributing to hyporeflexia during spinal shock, while it is diminished in chronic SCI, contributing to hyperreflexia associated with spasticity. These studies laid the foundation for investigating complete H recruitment curves to better understand neurophysiological behaviour. Hilgervood [19] examined M-wave characteristics and H-reflex thresholds with and without vibration, finding that vibration lowered H-reflex thresholds but did not affect the maximum H-reflex thresholds, and the M-wave remains consistent between subjects and conditions. Reduction in H-reflex due to vibration is attributed to presynaptic inhibition and post-activation depression. Murilo [23] extended vibration research to other muscle groups and with longer exposure time, applying 10 minutes of FMV to the quadriceps. This led to a decrease in the Soleus H/M ratio, with more prominent effects in complete SCIs. Vibration also reduced clonus frequency and duration and decreased the T-Reflex. The heteronymous H-reflex response in complete and incomplete SCIs was attributed to mechanical spread of vibration and the ‘busy line’ phenomenon, making fibres unresponsive to other inputs during vibration. Perez [20] examined the effects of focal vibration on the TA tendon in chronic complete SCIs, highlighting the H-reflex behaviour in reciprocal Ia and presynaptic D1 inhibition mechanisms in the Soleus muscle. Vibration inhibited H-reflex for reciprocal Ia inhibition (maintained up to 5 minutes) but did not have significant short or long-term effects on presynaptic D1 inhibition. The presynaptic Ia terminal and Ia interneuron were considered the possible sites of inhibition and presynaptic inhibition, post activation depression and robust spindle activation were attributed as the possible causes. Spasms are a common consequence after SCI. Butler [21] found that involuntary spasm-like EMG activity, evoked by superficial nerve stimulation in complete chronic SCIs, was reduced by vibrating Achilles tendon. The EMG activity depression was prominent in muscles proximal to the site of vibration but not in the distant ones. The vibration however did not affect the SOL H-reflex in SCIs. Persistent inward currents (PICs) were attributed to the long-lasting depression of the involuntary activity and not presynaptic terminal inhibition or motoneuron excitability. Cutaneous reflex responses were also explored. Gomez-Sariano [26] showed that vibration inhibited the long latency TA cutaneous reflex during plantarflexions in SCIs with spasticity, with the extent of inhibition correlated to the Modified Ashworth Scale (MAS), indicating greater reflex inhibition in subjects with higher spasticity. In contrast, DeForest [28] noted that tendon vibration at specific frequencies inhibited the long-lasting component of the cutaneous reflex in antagonist muscles but not in agonist muscles, offering insights into the suppression of PICs and the activation of interneurons involved in central pattern generator networks.

In conclusion, these studies contribute significantly to our understanding of neural mechanisms affected by FMV in SCIs. They highlight the potential for vibration to induce plastic changes in neural circuitry and improve spasticity management. Future research should continue to explore the implications of different vibration frequencies, stimulation location (muscle Vs tendon, agonist Vs antagonist), with and without accompanying muscle contraction/activity and the potential for long-term neural modulation.

## 5. CONCLUSION

In conclusion, the findings presented in this systematic review shed light on the use of FMV as a potential therapeutic approach for improving mobility and strength and understanding sensorimotor function in individuals with SCIs. The reviewed studies encompassed a total of 422 SCI patients and 132 non-disabled participants, with a predominant focus on male individuals and chronic SCI cases. However, a significant limitation was the lack of specification regarding the completeness of the SCI, which may affect the interpretation of results. Geographically, most of the studies were conducted in North America, highlighting regional disparities in research priorities and resource availability. Another limitation is the cohort sizes of the available studied, as none of the studies were powered studies. Several early studies had a sample size approaching 60 but did not record if the sample size was powered or not, the remaining were convenience samples. The smaller sample sizes reduce the confidence in studies’ conclusion to be extrapolated.

FMV interventions primarily used frequencies in the 60-80 Hz range, suggesting an established optimal range. The exploration of FMV in the context of SCI rehabilitation yields a complex yet promising narrative. It appears that FMV can have both short-term and long-term effects on sensory and motor function, mobility, and muscle strength. However, the specific outcomes and their sustainability may vary, highlighting the need for further research and clinical studies to uncover the underlying mechanisms and determine the most effective strategies for incorporating FMV into SCI rehabilitation programs. As discussed in the introduction section, neurophysiological mechanisms underpinning the effects of FMV in healthy are relatively well studied. With modulating effects of FMV on a) muscle afferents [38, 39, 40], b) cutaneous mechanoreceptors [46], c) cortical excitability [47, 48] and d) TVR[38] well documented in healthy. However, with the exception of one study [28], the underlying mechanisms in response to FMV in SCI population have not been investigated. This gap in our understanding not only limits the translation of FMV’s effects observed in healthy to SCI rehabilitation, it also raises questions about persistence of the above observed effects longitudinally to positively effect rehabilitation outcome measures.

On the positive side, findings reported and discussed in this review do provide a snapshot of ongoing efforts to improve the quality of life for individuals with SCI through innovative rehabilitation techniques like FMV. The ideal parameters for FMV, however, require further investigation. Furthermore, different vibration devices are known to have intra- and inter-device variability [50], which translates to difference in their performance and output effects. A possibility of variation in the vibration parameters and their consequent performance should hence be considered.

## 6. STUDY LIMITATIONS

This systematic review has certain limitations to consider. The included studies exhibited substantial heterogeneity in design, FMV parameters, and participant characteristics, making direct comparisons and generalizability challenging. Small sample sizes, limited diversity in SCI severity and duration, and the absence of control groups in some studies raise questions about the robustness of the findings. Variation in vibration devices is another question. Regional bias and potential publication bias may affect the generalizability of results. Many studies focused on short-term outcomes, while long-term effects and underlying mechanisms were often underexplored. These limitations emphasize the need for more standardized, diverse, and mechanistic research to better understand the potential and practical application of FMV in SCI rehabilitation.

## Data Availability

All data produced in the present work are contained in the manuscript

## REFERENCES

[1] Nas, K. (2015). Rehabilitation of spinal cord injuries. World Journal of Orthopedics, 6(1), 8. doi:10.5312/wjo.v6.i1.8

[2] Simpson, L. A., Eng, J. J., Hsieh, J. T. C., & Wolfe and the Spinal Cord Injury Re, D. L. (2012). The health and life priorities of individuals with Spinal Cord Injury: A systematic review. Journal of Neurotrauma, 29(8), 1548–1555. doi:10.1089/neu.2011.2226

[3] Lanig, I. S., New, P. W., Burns, A. S., Bilsky, G., Benito-Penalva, J., Bensmail, D., & Yochelson, M. (2018). Optimizing the management of spasticity in people with spinal cord damage: A clinical care pathway for assessment and treatment decision making from the Ability Network, an international initiative. Archives of Physical Medicine and Rehabilitation, 99(8), 1681–1687. doi:10.1016/j.apmr.2018.01.017

[4] Gohritz, A., & Fridén, J. (2018). Management of spinal cord injury-induced upper extremity spasticity. Hand Clinics, 34(4), 555–565. doi:10.1016/j.hcl.2018.07.001

[5] Sadeghi, M., & Sawatzky, B. (2014). Effects of vibration on spasticity in individuals with Spinal Cord Injury. American Journal of Physical Medicine & amp; Rehabilitation, 93(11), 995–1007. doi:10.1097/phm.0000000000000098

[6] Herszkowicz, I., Beric, A., & Lindblom, U. (1986). Vibratory perception thresholds at the clavicle in patients with spinal cord injury. Journal of Neurology, Neurosurgery & Psychiatry, 49(9), 1063–1065. doi:10.1136/jnnp.49.9.1063

[7] Takakura, N., Iijima, S., Kanamaru, A., Shibuya, M., Homma, I., & Ohashi, M. (1996). Vibration-induced finger flexion reflex and inhibitory effect of acupuncture on this reflex in cervical spinal cord injury patients. Neuroscience Research, 26(4), 391–394. doi:10.1016/s0168-0102(96)01119-4

[8] Ribot-Ciscar, E., Butler, J. E., & Thomas, C. K. (2003). Facilitation of triceps brachii muscle contraction by tendon vibration after chronic cervical spinal cord injury. Journal of Applied Physiology, 94(6), 2358–2367. doi:10.1152/japplphysiol.00894.2002

[9] Backus, D., Cordo, P., Gillott, A., Kandilakis, C., Mori, M., & Raslan, A. M. (2014). Assisted Movement with proprioceptive stimulation reduces impairment and restores function in incomplete spinal cord injury. Archives of Physical Medicine and Rehabilitation, 95(8), 1447–1453. doi:10.1016/j.apmr.2014.03.011

[10] Gomes-Osman, J., & Field-Fote, E. C. (2014). Cortical vs. afferent stimulation as an adjunct to functional task practice training: A randomized, comparative pilot study in people with cervical spinal cord injury. Clinical Rehabilitation, 29(8), 771–782. doi:10.1177/0269215514556087

[11] Etoom, M., & Marchetti, A. (2015). Effect of a focal muscle vibration above triceps brachii muscle on upper limb spasticity in a patient with a chronic spinal cord injury: A case report. International Journal of Physiotherapy and Research, 3(4), 1171–1174. doi:10.16965/ijpr.2015.162

[12] Fusco, G., Tidoni, E., Barone, N., Pilati, C., & Aglioti, S. M. (2016). Illusion of arm movement evoked by tendon vibration in patients with spinal cord injury. Restorative Neurology and Neuroscience, 34(5), 815–826. doi:10.3233/rnn-160660

[13] Vojinovic, T. J., Linley, E., Zivanovic, A., & Rui Loureiro, C. V. (2019). Effects of focal vibration and robotic assistive therapy on upper limb spasticity in incomplete spinal cord injury. 2019 IEEE 16th International Conference on Rehabilitation Robotics (ICORR). doi:10.1109/icorr.2019.8779566

[14] Tazoe, T., & Perez, M. A. (2021). Abnormal changes in motor cortical maps in humans with spinal cord injury. The Journal of Physiology, 599(22), 5031–5045. doi:10.1113/jp281430

[15] Ashby, P., Verrier, M., & Lightfoot, E. (1974). Segmental reflex pathways in spinal shock and spinal spasticity in man. Journal of Neurology, Neurosurgery & Psychiatry, 37(12), 1352–1360. doi:10.1136/jnnp.37.12.1352

[16] Verrier, M., MacLeod, S., & Ashby, P. (1975). The effect of diazepam on presynaptic inhibition in patients with complete and incomplete spinal cord lesions. Canadian Journal of Neurological Sciences / Journal Canadien Des Sciences Neurologiques, 2(3), 179–184. doi:10.1017/s0317167100020229

[17] Taylor, S., Ashby, P., & Verrier, M. (1984). Neurophysiological changes following traumatic spinal lesions in man. Journal of Neurology, Neurosurgery & Psychiatry, 47(10), 1102–1108. doi:10.1136/jnnp.47.10.1102

[18] Calancie, B., Broton, J. G., John Klose, K., Traad, M., Difini, J., & Ram Ayyar, D. (1993). Evidence that alterations in presynaptic inhibition contribute to segmental hypo- and hyperexcitability after spinal cord injury in man. Electroencephalography and Clinical Neurophysiology/Evoked Potentials Section, 89(3), 177–186. doi:10.1016/0168-5597(93)90131-8

[19] Hilgevoord, A. A. J., Koelman, J. H. T. M., Bour, L. J., & de Visser, B. W. O. (1996). The relationship between the soleus H-reflex amplitude and vibratory inhibition in controls and spastic subjects. I. Experimental results. Journal of Electromyography and Kinesiology, 6(4), 253–258. doi:10.1016/s1050-6411(96)00006-5

[20] Perez, M. A., Floeter, M. K., & Pield-Fote, E. (2004). Repetitive sensory input increases reciprocal IA inhibition in individuals with incomplete spinal cord injury. Journal of Neurologic Physical Therapy, 28(3), 114–121. doi:10.1097/01253086-200409000-00003

[21] Butler, J. E., Godfrey, S., & Thomas, C. K. (2006). Depression of involuntary activity in muscles paralyzed by spinal cord injury. Muscle & Nerve, 33(5), 637–644. doi:10.1002/mus.20500

[22] Cotey, D., George Hornby, T., Gordon, K. E., & Schmit, B. D. (2009). Increases in muscle activity produced by vibration of the thigh muscles during locomotion in chronic human spinal cord injury. Experimental Brain Research, 196(3), 361–374. doi:10.1007/s00221-009-1855-9

[23] Murillo, N., Kumru, H., Vidal-Samso, J., Benito, J., Medina, J., Navarro, X., & Valls-Sole, J. (2011). Decrease of spasticity with muscle vibration in patients with spinal cord injury. Clinical Neurophysiology, 122(6), 1183–1189. doi:10.1016/j.clinph.2010.11.012

[24] Field-Fote, E., Ness, L. L., & Ionno, M. (2012). Vibration elicits involuntary, step-like behavior in individuals with Spinal Cord Injury. Neurorehabilitation and Neural Repair, 26(7), 861–869. doi:10.1177/1545968311433603

[25] Onushko, T., Hyngstrom, A., & Schmit, B. D. (2013). Hip proprioceptors preferentially modulate reflexes of the leg in human spinal cord injury. Journal of Neurophysiology, 110(2), 297–306. doi:10.1152/jn.00261.2012

[26] Gómez-Soriano, J., Serrano-Muñoz, D., Bravo-Esteban, E., Avendaño-Coy, J., Ávila-Martin, G., Galán-Arriero, I., & Taylor, J. (2018). Afferent stimulation inhibits abnormal cutaneous reflex activity in patients with spinal cord injury spasticity syndrome. NeuroRehabilitation, 43(2), 135–146. doi:10.3233/nre-172404

[27] Bochkezanian, V., Newton, R. U., Trajano, G. S., Vieira, A., Pulverenti, T. S., & Blazevich, A. J. (2018). Effect of tendon vibration during wide-pulse neuromuscular electrical stimulation (NMES) on muscle force production in people with Spinal Cord Injury (SCI). BMC Neurology, 18(1). doi:10.1186/s12883-018-1020-9

[28] DeForest, B. A., Bohorquez, J., & Perez, M. A. (2020). Vibration attenuates spasm-like activity in humans with Spinal Cord Injury. The Journal of Physiology, 598(13), 2703–2717. doi:10.1113/jp279478

[29] Li, Y., Wei, B., Zhong, Y. et al. A bibliometric analysis of global research on spinal cord injury: 1999–2019. Spinal Cord 60, 281–287 (2022). 10.1038/s41393-021-00691-9

[30] McDaid D, Park AL, Gall A, Purcell M, Bacon M. Understanding and modelling the economic impact of spinal cord injuries in the United Kingdom. Spinal Cord. 2019 Sep;57(9):778-788. doi: 10.1038/s41393-019-0285-1. Epub 2019 May 13. PMID: 31086273; PMCID: PMC6760568.

[31] Merritt CH, Taylor MA, Yelton CJ, Ray SK. Economic impact of traumatic spinal cord injuries in the United States. Neuroimmunol Neuroinflamm. 2019;6:9. doi: 10.20517/2347-8659.2019.15. Epub 2019 Jul 20. PMID: 33869674; PMCID: PMC8052100.

[32] Islam, M.S., Harvey, L.A., Hossain, M.S. et al. The cost of providing a communitybased model of care to people with spinal cord injury, and the healthcare costs and economic burden to households of spinal cord injury in Bangladesh. Spinal Cord 59, 833–841 (2021). 10.1038/s41393-020-00600-6

[33] Peter Francis Raguindin, Taulant Muka, Marija Glisic, Sex and gender gap in spinal cord injury research: Focus on cardiometabolic diseases. A mini review, Maturitas, Volume 147, 2021, Pages 14-18, ISSN 0378-5122, 10.1016/j.maturitas.2021.03.004.

[34] Elbasiouny SM, Moroz D, Bakr MM, Mushahwar VK. Management of Spasticity After Spinal Cord Injury: Current Techniques and Future Directions. Neurorehabilitation and Neural Repair. 2010;24(1):23-33. doi:10.1177/1545968309343213

[35] Fattorini, L.; Rodio, A.; Filippi, G.M.; Pettorossi, V.E. Effectiveness of Focal Muscle Vibration in the Recovery of Neuromotor Hypofunction: A Systematic Review. J. Funct. Morphol. Kinesiol. 2023, 8, 103. 10.3390/jfmk8030103

[36] Moggio L, de Sire A, Marotta N, Demeco A, Ammendolia A. Vibration therapy role in neurological diseases rehabilitation: an umbrella review of systematic reviews. Disabil Rehabil. 2022 Oct;44(20):5741-5749. doi: 10.1080/09638288.2021.1946175. Epub 2021 Jul 5. PMID: 34225557.

[37] Fattorini, L.; Rodio, A.; Pettorossi, V.E.; Filippi, G.M. Is the Focal Muscle Vibration an Effective Motor Conditioning Intervention? A Systematic Review. J. Funct. Morphol. Kinesiol. 2021, 6, 39. 10.3390/jfmk6020039

[38] G. Eklund, K.-E. Hagbarth, Normal variability of tonic vibration reflexes in man, Experimental Neurology, Volume 16, Issue 1, 1966, Pages 80-92, ISSN 0014-4886, 10.1016/0014-4886(66)90088-4.

[39] Beverly Bishop, Vibratory Stimulation: Part I. Neurophysiology of Motor Responses Evoked by Vibratory Stimulation, Physical Therapy, Volume 54, Issue 12, December 1974, Pages 1273–1282, 10.1093/ptj/54.12.1273

[40] Burke, D, Hagbarth, K E, Löfstedt, L, Wallin, B G, (1976), The responses of human muscle spindle endings to vibration during isometric contraction.. The Journal of Physiology, 261 doi: 10.1113/jphysiol.1976.sp011581.

[41] Marconi B, Filippi GM, Koch G, et al. Long-Term Effects on Cortical Excitability and Motor Recovery Induced by Repeated Muscle Vibration in Chronic Stroke Patients. Neurorehabilitation and Neural Repair. 2011;25(1):48-60. doi:10.1177/1545968310376757

[42] Cordo P, Wolf S, Rymer WZ, Byl N, Stanek K, Hayes JR. Assisted Movement With Proprioceptive Stimulation Augments Recovery From Moderate-To-Severe Upper Limb Impairment During Subacute Stroke Period: A Randomized Clinical Trial. Neurorehabilitation and Neural Repair. 2022;36(3):239-250. doi:10.1177/15459683211063159

[43] Conrad MO, Scheidt RA, Schmit BD. Effects of Wrist Tendon Vibration on Targeted Upper-Arm Movements in Poststroke Hemiparesis. Neurorehabilitation and Neural Repair. 2011;25(1):61-70. doi:10.1177/1545968310378507

[44] Avvantaggiato C, Casale R, Cinone N, Facciorusso S, Turitto A, Stuppiello L, Picelli A, Ranieri M, Intiso D, Fiore P, Ciritella C, Santamato A. Localized muscle vibration in the treatment of motor impairment and spasticity in post-stroke patients: a systematic review. Eur J Phys Rehabil Med. 2021 Feb;57(1):44-60. doi: 10.23736/S1973-9087.20.06390-X. Epub 2020 Oct 28. PMID: 33111513.

[45] Celletti C, Suppa A, Bianchini E, Lakin S, Toscano M, La Torre G, Di Piero V, Camerota F. Promoting post-stroke recovery through focal or whole body vibration: criticisms and prospects from a narrative review. Neurol Sci. 2020 Jan;41(1):11-24. doi: 10.1007/s10072-019-04047-3. Epub 2019 Aug 30. PMID: 31468237.

[46] Freeman, Alan W., Johnson, Kenneth O., (1982), Cutaneous mechanoreceptors in macaque monkey: temporal discharge patterns evoked by vibration, and a receptor model. The Journal of Physiology, 323 doi: 10.1113/jphysiol.1982.sp014059.

[47] Thomas F. Münte, E. Michael Jöbges, Bernardina M. Wieringa, Sebastian Klein, Margot Schubert, Sönke Johannes, Reinhard Dengler, Human evoked potentials to long duration vibratory stimuli: role of muscle afferents, Neuroscience Letters, Volume 216, Issue 3, 1996, Pages 163–166, ISSN 0304-3940, 10.1016/0304-3940(96)13036-6.

[48] Barss TS, Collins DF, Miller D and Pujari AN (2021) Indirect Vibration of the Upper Limbs Alters Transmission Along Spinal but Not Corticospinal Pathways. Front. Hum. Neurosci. 15:617669. doi: 10.3389/fnhum.2021.617669

[49] Page M J, Moher D, Bossuyt P M, Boutron I, Hoffmann T C, Mulrow C D et al. PRISMA 2020 explanation and elaboration: updated guidance and exemplars for reporting systematic reviews BMJ 2021; 372 :160 doi:10.1136/bmj.n160

[50] Botter A, Cerone GL, Saggini R, Massazza G, Minetto MA. Characterization of the stimulation output of four devices for focal muscle vibration. Med Eng Phys. 2020 Nov;85:97-103. doi: 10.1016/j.medengphy.2020.10.002. Epub 2020 Oct 7. PMID: 33081969.

